# Target Trial Emulation of Metformin for Dementia Prevention in All of Us: Robustness to Cohort and Confounder Definitions

**DOI:** 10.64898/2026.11.02.26367055

**Authors:** Marie-Laure Charpignon, Tal Shnitzer, Aleena Iraqui, Danielle Pace, Mahnaz Maddah, Caroline Uhler

## Abstract

Metformin use has been associated with a lower risk of incident dementia than other antidia-betics. We perform target trial emulation and causal competing risk analysis in the NIH All of Us (AoU) Research Program database, contrasting the effects of metformin and sulfony-lurea initiation on dementia onset and death without dementia. AoU’s social and behavioral health data allowed us to test robustness to excluding such confounders, and we further adjusted for calendar year effects and prior antidiabetic exposure, using both semiparametric Cox proportional hazards (PH) models and nonparametric models. Under stabilized inverse propensity score of treatment weighting, metformin was associated with a substantially lower risk of death (hazard ratio [HR]: 0.61, 95% CI: [0.49, 0.75]) and a lower incidence of dementia (HR: 0.90, 95% CI: [0.82, 1.02]). Estimated HRs were closely aligned with estimates obtained in the UK primary care and US Mass General Brigham health systems. This finding was robust to the inclusion of participants without a type 2 diabetes diagnosis at baseline and to the consideration of different covariate sets in the propensity score model, supporting metformin as a promising candidate for dementia prevention and motivating prospective validation.

**Data and Code Availability:** This study used data from the NIH All of Us (AoU) Research Program’s Controlled Tier Dataset (CDRv9), available to authorized users on the Researcher Workbench. Analysis code is attached as supplemental material.

## 1. Introduction

Drug repurposing offers a faster and cheaper route to therapy than de-novo development. The growing availability of electronic health records (EHRs) enables analysis of drug exposure and disease trajectories in populations that may be more representative than trial participants. In this context, the target trial emulation (TTE) framework (Hernan and Robins, 2016) provides a powerful tool to design insilico trials that combine the rigor of a randomized controlled trial (RCT) with the long follow-up available in observational studies.

Dementia has no disease-modifying treatment, motivating the repurposing of existing drugs. To date, 45% of dementia risk has been attributed to modifiable risk factors, including type 2 diabetes (T2D) (Chatterjee et al., 2016; Livingston et al., 2024). Metformin, the first-line therapy for T2D, has been associated with extended lifetime and lower dementia incidence than placebo or other antidiabetics in observational studies and animal models. Recently, its use has been extended beyond T2D, including preventatively in patients diagnosed with prediabetes. A pilot RCT of short-acting metformin provided initial evidence of its efficacy in slowing cognitive decline in people with mild cognitive impairment without T2D (Luchsinger et al., 2016). Whether these cognitive benefits hold in routine care data remains unclear.

Charpignon et al. (2022) emulated a target trial to interrogate the effect of initiating metformin vs. an antidiabetic from the sulfonylurea drug class on dementia onset and competing death, using data from patients in the UK primary care and US Mass General Brigham (MGB) health systems. A protective association was found for both outcomes. However, important confounders such as lifestyle and socioeconomic factors were unmeasured and could not be taken into account. Moreover, the representation of Black and Hispanic people was limited, and the ten-year study period excluded the COVID-19 pandemic.

We replicated this study design in the AoU Research Program, a large and diverse US cohort with rich survey data, social and behavioral information, a long follow-up allowing the evaluation of calendar year effects, and a sufficient sample size to stratify by T2D status. Following the inverse propensity score of treatment weighting approach in Charpignon et al. (2022), we conducted causal analyses to investigate these aspects by restricting the population to participants with a T2D diagnosis at baseline and by enriching the propensity score model with calendar year of first prescription, lifestyle, education and socioeconomic variables, as well as prior history of other antidiabetic drug prescriptions. We aim to provide insight into the potential benefits of metformin in the general adult population, and to assess the extent of balance between arms at baseline with respect to social and behavioral health factors often left unmeasured in prior studies, quantify their incremental contribution to confounding control, and evaluate the robustness of treatment effect estimates.

## 2. Methods

### Design and data

We emulated a target trial in AoU, comparing initiation of metformin vs. sulfonylureas on time to dementia onset, with death without dementia as a competing event. We adapted the target trial emulation design of Charpignon et al. (2022), reusing their causalCmprsk estimator while refining eligibility criteria and extending the confounder set.

### Cohort

Eligible participants were adults age *≥*50 at their first metformin or sulfonylurea prescription, defined as baseline. At baseline, participants were required to have available sex at birth information, antidiabetic monotherapy, and no dementia diagnosis (Figure 1). Participants with chronic kidney disease (CKD) at baseline were excluded from both treatment arms because CKD may preclude metformin use (Imam, 2017) and therefore could determine treatment assignment. Applying this restriction also improved comparability of baseline kidney function between treatment arms. We further restricted the cohort to include only participants with a T2D diagnosis at baseline, to obtain a more homogeneous cohort. Arms were defined by the first prescription drug. Drugs were mapped using ATC code, while dementia and comorbidities were defined from curated ICD/SNOMED code sets and drug lists when appropriate. These appear in CSV files with the code.

**Figure 1:**
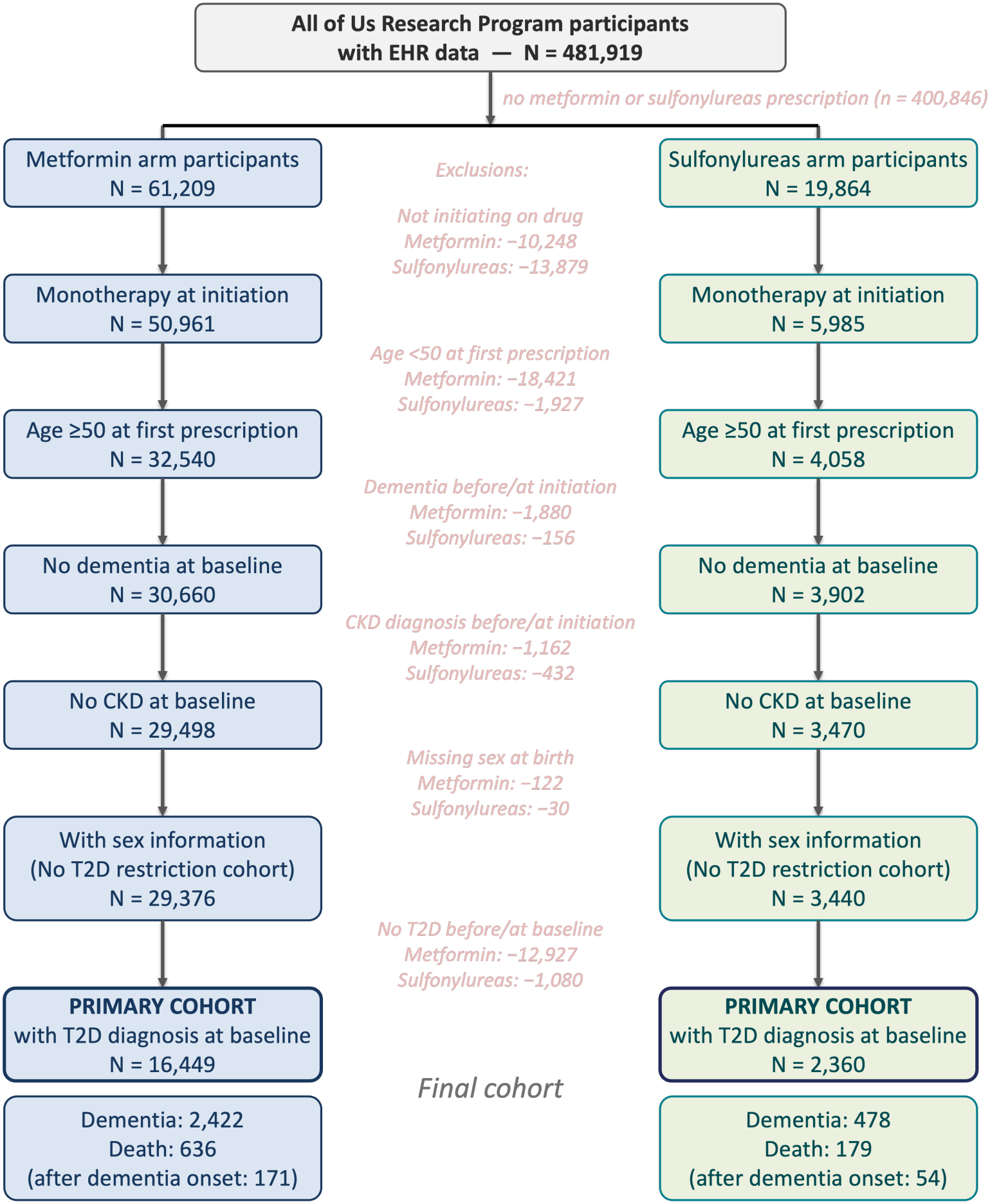
CONSORT diagram: cohort filtering and participant counts. Boxes show the number of participants remaining in each arm after each eligibility criterion is applied; red text between the arms gives the per-arm exclusion counts at each step.

### Confounders

As in Charpignon et al. (2022), the propensity score model included age at first prescription, sex at birth, race, ethnicity, baseline comorbidities (stroke, hypertension, obesity, CVD, cancer), and binned BMI and HbA1c, measured up to three years before baseline. Missing values were kept as explicit categories. We additionally included calendar year of first prescription, binned into groups of 3-5 years to alleviate prescribing shifts, lifestyle factors (smoking and alcohol use), education information from AoU surveys, neighborhood-level zipcode-linked socioeconomic measures (Appendix B), and prior exposure to other antidiabetic drug classes (DPP-4 inhibitors, SGLT2 inhibitors, GLP-1 receptor agonists, insulin) since participants are not required to be treatmentnaive and prior use may proxy for diabetes severity or partial treatment history. The continuous covariates in the propensity model were standardized to zero mean and unit variance, and neighborhood-level measures were median-imputed.

### Effect estimation

Baseline randomization was emulated via inverse propensity score of treatment weighting. Logistic regression was used to estimate propensity scores. Our main analysis focused on the average treatment effect (ATE) as the causal estimand of interest and used stabilized ATE weights. Our secondary analysis considered the average treatment effect in the overlap population (ATO) as the estimand and involved overlap weights. Covariate balance before vs. after weighting was assessed via absolute standardized mean differences (threshold 0.1, following the literature’s convention). Under each weighting scheme, we estimated hazard ratios (HRs) and cumulative risks of dementia onset and death without dementia, both nonparametrically and via cause-specific Cox PH models, using causalCmprsk with a Bayesian bootstrap for uncertainty estimation (1,000 replicates). For the two competing events, we additionally derived time-varying risk differences and the restricted mean time lost or gained due to treatment, following the definitions in Vakulenko-Lagun et al. (2023); Charpignon et al. (2022).

### Robustness scenarios

Beyond the primary analysis, we performed five sensitivity analyses: (i) the choice of balancing weights (stabilized ATE vs. overlap); (ii) incremental contribution of calendar year of first prescription; (iii) incremental contribution of lifestyle covariates; (iv) incremental contribution of educational attainment and neighborhood characteristics information; and (v) including all initiators regardless of a coded T2D diagnosis, while adding it as a comorbidity. Consideration (v) highlights an improvement over Charpignon et al. (2022), which did not distinguish patients with a coded T2D diagnosis from others, resulting in the potential mixture of populations with distinct indications. Indeed, additional clinical uses of metformin have emerged beyond T2D, including polycystic ovarian syndrome, prevention in prediabetes, adjunct therapy in type 1 diabetes, and healthy aging (Petrie, 2024; Srivastava, 2024).

## 3. Results

### Cohort

After excluding ineligible participants, our cohort comprised *N* =18,809 participants (metformin: *N* =16,449, sulfonylureas: *N* =2,360). During follow-up, 2,900 dementia events (metformin: 2,422, sulfonylureas: 478) and 815 death events were recorded, including 225 deaths after dementia onset (metformin: 636 and 171, sulfonylureas: 179 and 54).

Distributions of baseline characteristics stratified by treatment arm appear in Tables 2 and 3. Age and neighborhood-level measures were similar across arms, but HbA1c, obesity, alcohol use, and prior antidiabetic exposure showed significant differences. Sulfonylureas initiators were less likely to have HbA1c *<* 7% (20.3% vs. 33.2%), less likely to have a diagnosis of obesity (12.1% vs. 17.8%), more likely to report no alcohol use (42.8% vs. 37.0%), and more likely to have prior insulin exposure (22.2% vs. 19.7%).

**Table 1:** Hazard ratios (metformin vs. sulfonylureas) for dementia and death in Cox PH models with alternative weighting schemes, in the T2D-restricted cohort. HR [95% CI].

| **Weighting** | **Outcome** | **Main** |
|---|---|---|
| Unadjusted | Dementia | 1.01 [0.91, 1.12] |
|  | Death | 0.82 [0.67, 1.00] |
| Overlap | Dementia | 0.92 [0.84, 1.02] |
|  | Death | 0.67 [0.55, 0.81] |
| Stabilized ATE | Dementia | 0.90 [0.82, 1.02] |
|  | Death | 0.61 [0.49, 0.75] |

**Table 2:**
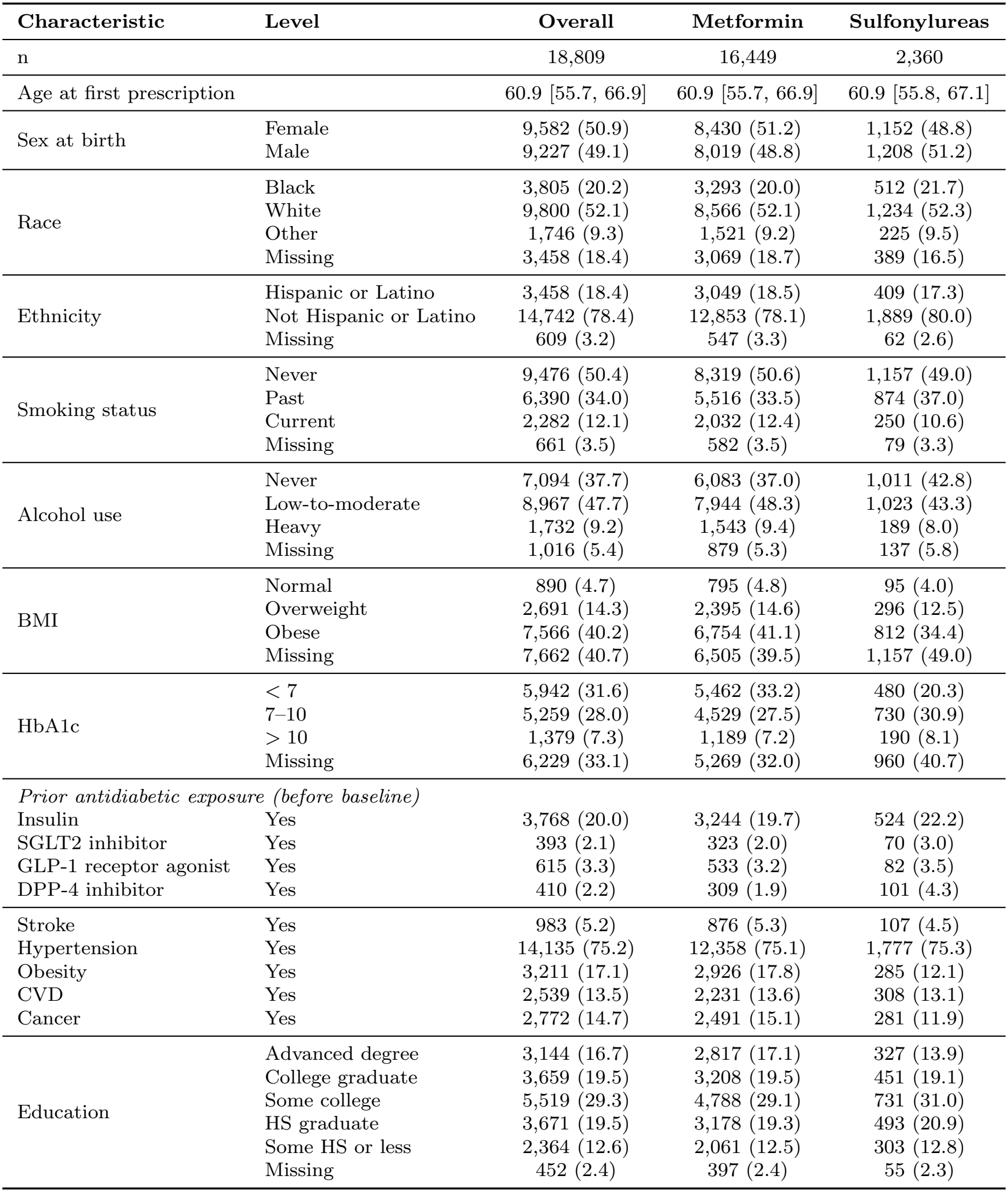
Baseline characteristics of metformin and sulfonylurea initiators with a T2D diagnosis at baseline (primary cohort). Continuous variables: median [interquartile range], categorical variables, n (%).

**Table 3:** Baseline characteristics of metformin and sulfonylurea initiators: neighborhood-level variables derived from survey linkages using residential zipcodes. The median is provided, along with the interquartile range. Out of *N* = 18,809 participants, 844 had missing neighborhood-level data.

| **Characteristic** | **Overall** | **Metformin** | **Sulfonylureas** |
|---|---|---|---|
| Assisted income, % of zipcode | 13.3 [10.7, 16.8] | 13.3 [10.7, 16.7] | 13.3 [11.0, 17.1] |
| High school education, % of zipcode | 88.3 [84.7, 91.5] | 88.3 [84.7, 91.5] | 88.1 [84.7, 91.5] |
| Median household income, \$ | 61,390 [55,434, 73,170] | 61,390 [55,523, 73,838] | 61,024 [54,647, 73,170] |
| No health insurance, % of zipcode | 8.6 [5.5, 11.5] | 8.6 [5.5, 11.5] | 9.2 [5.7, 11.5] |
| Poverty, % of zipcode | 14.9 [11.8, 18.2] | 14.9 [11.8, 18.2] | 14.9 [12.1, 18.7] |

### Propensity scores

The estimated propensity score distribution shifted to the right, but substantial overlap remained (Figure 2). Both weighting schemes reduced absolute standardized mean differences below the 0.1 threshold, with overlap weights driving them to zero by construction (Figure 3).

**Figure 2:**
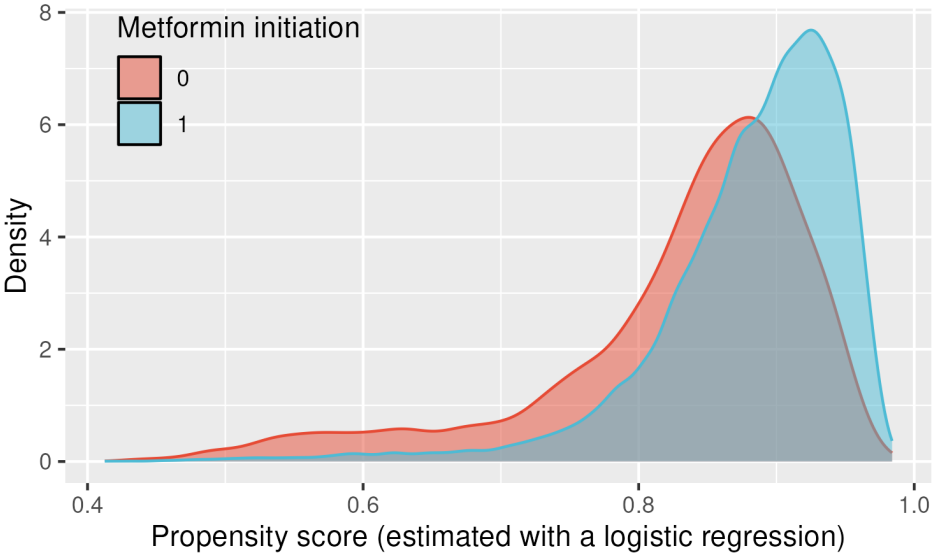
Estimated propensity score density by treatment arm (metformin = 1, sulfonylurea = 0), main analysis.

**Figure 3:**
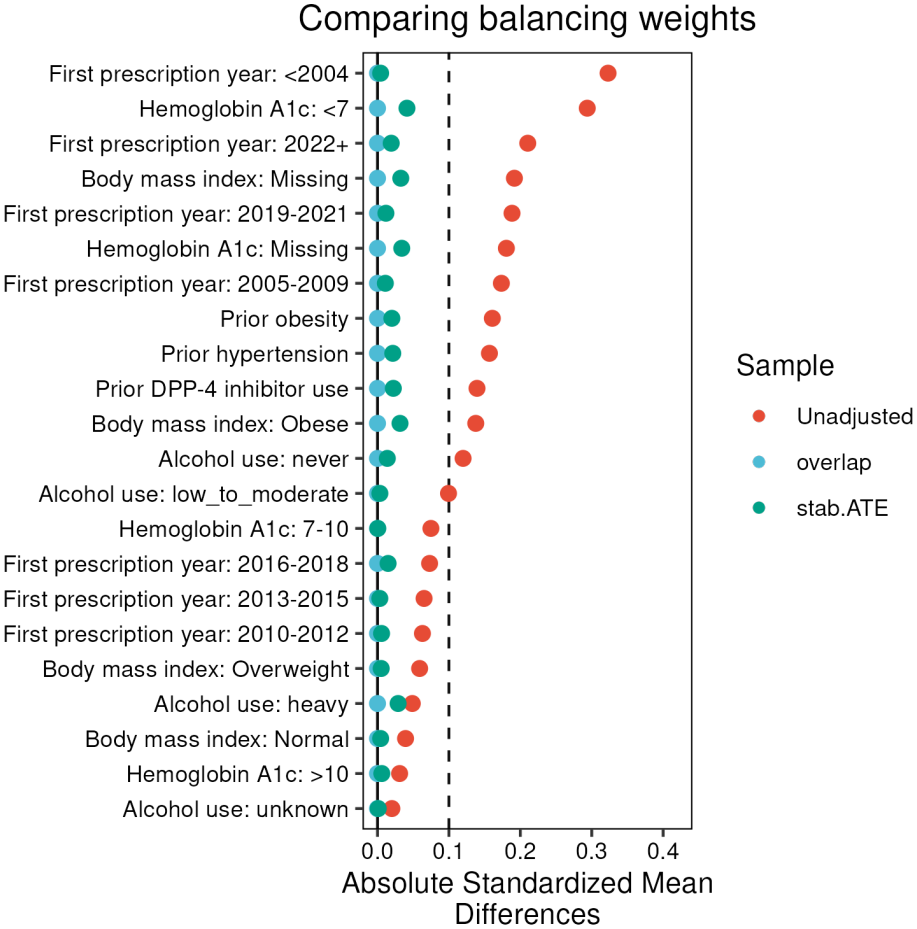
Absolute standardized mean differences for the covariates with the largest imbalance, by weighting scheme. Dashed line: 0.1.

### Treatment effect estimates

As shown in Table 1, both weighting schemes shifted the estimated HR for dementia onset toward a similar protective effect (overlap: 0.92, [0.84, 1.02], stabilized ATE: 0.90, [0.82, 1.02]). For competing death, metformin was protective under all weighting schemes (overlap: 0.67, [0.55, 0.81], stabilized ATE: 0.61, [0.49, 0.75]). Compared to Charpignon et al. (2022) (dementia HR 0.81–0.86, death HR 0.57–0.66), the signal obtained for dementia onset is consistent but attenuated, while the competing death benefit is reproduced.

For both outcomes and all three weighting schemes, the nonparametric log HR curve tracked the constant HR within its 95% CI band through 15 years of follow-up (Figures 4, 5 and 6), supporting the PH assumption. Some divergence appears beyond 15 years, in part because of a smaller number of participants.

**Figure 4:**
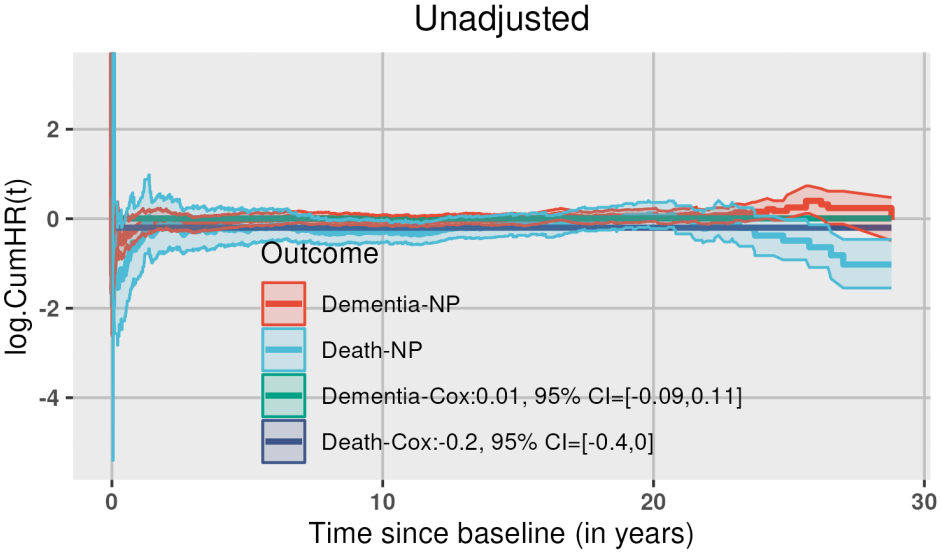
Log hazard ratios estimated via the Cox PH model vs. the nonparametric approach, for both dementia and death (unadjusted weights).

**Figure 5:**
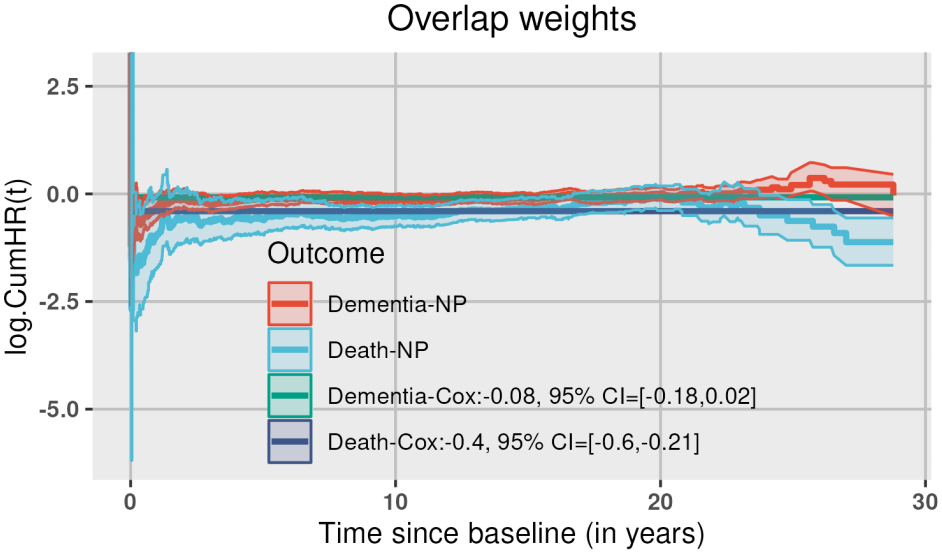
Log hazard ratios estimated via the Cox PH model vs. the nonparametric approach, for both dementia and death (over-lap weights).

**Figure 6:**
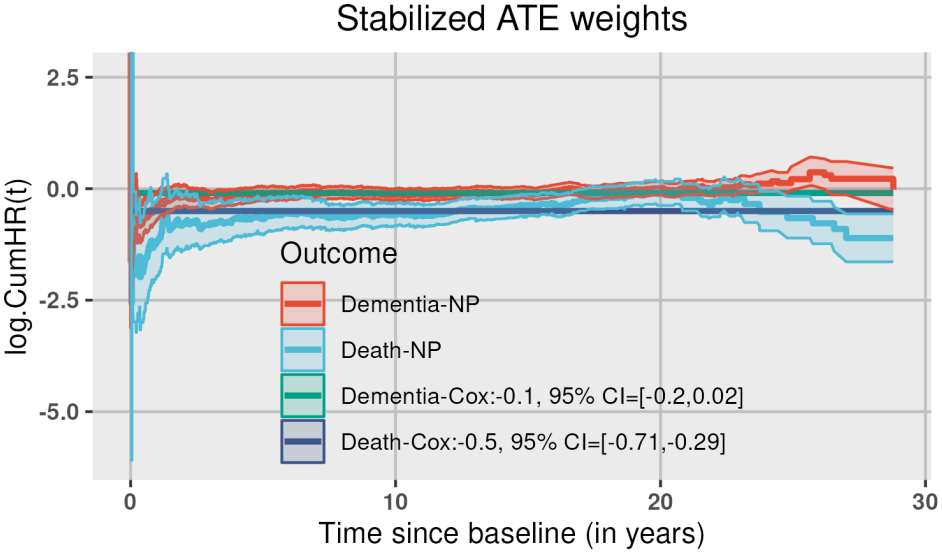
Log hazard ratios estimated via the Cox PH model vs. the nonparametric approach, for both dementia and death (stabilized ATE weights).

The cumulative risk of dementia was estimated at 28–30% within 15 years of baseline, with metformin tracking below sulfonylureas by about 2% (Figure 7). Throughout follow-up, the cumulative risk of death without dementia was substantially lower under metformin initiation than under sulfonylurea initiation (6% vs. 9% by year 15). The dementia risk difference (Figure 8) was negative (favoring metformin initiation), with confidence intervals spanning zero, whereas the death risk difference remained negative. The restricted mean time analysis (Figure 9) also favored metformin initiation. Within 15 years of baseline, *∼*75 dementia-free days and 96 days of life were gained on average with metformin initiation.

**Figure 7:**
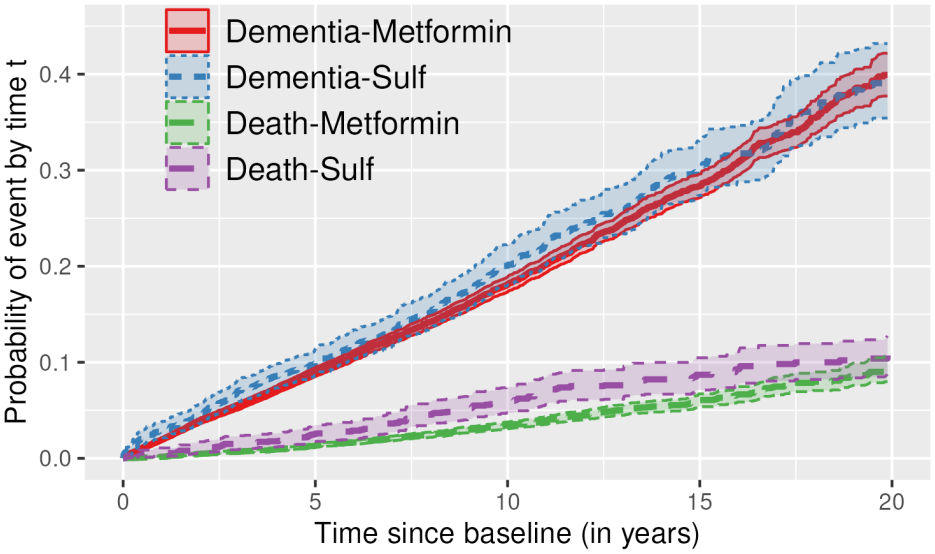
Cumulative incidence functions for dementia and competing death, by arm, estimated with the Cox model using stabilized ATE weighting.

**Figure 8:**
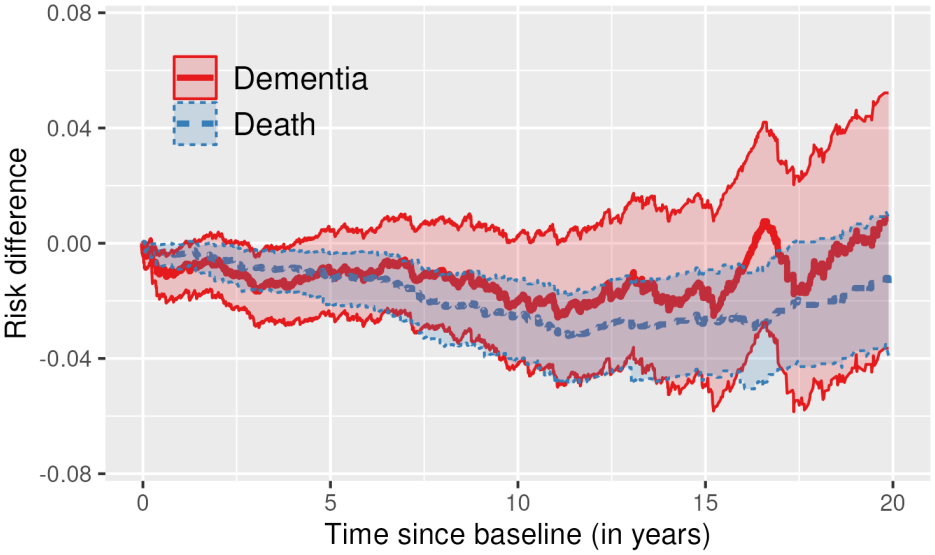
Time-varying risk difference curves (metformin *−* sulfonylureas) for dementia and competing death, estimated with the Cox model using stabilized ATE weighting. Negative risk difference values favor metformin.

**Figure 9:**
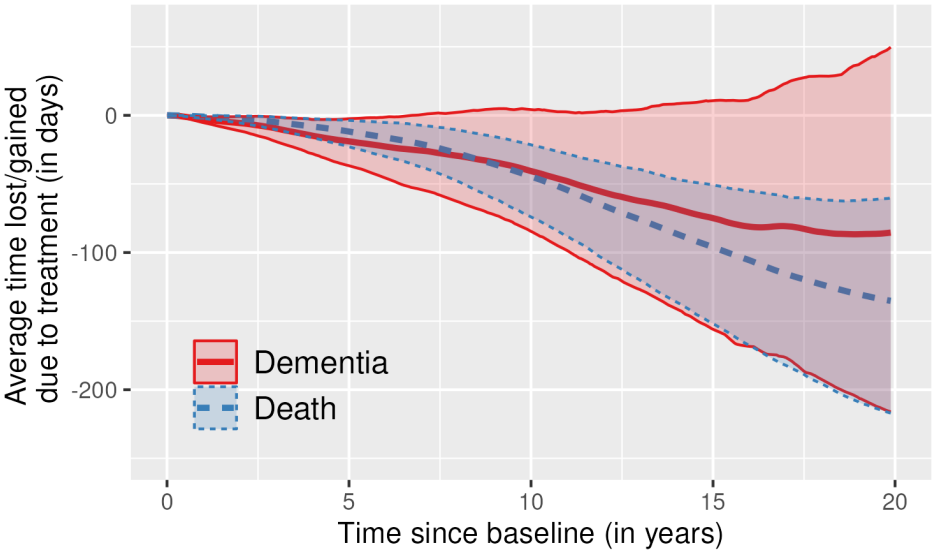
Restricted mean time lost/gained due to treatment (days), for dementia and competing death, estimated with the Cox model using stabilized ATE weighting. Negative values favor metformin

### Contribution of individual confounders

Enriching the propensity score model variably affected arm separation (Figure 10). The distribution of the calendar year of first prescription showed a marked temporal structure both overall volume and by treatment arm (Figure 11), consistent with clinical guidelines recommending metformin as the firstline antidiabetic therapy in 2006 (Nathan et al., 2006), pandemic-related disruptions to routine primary care, and the AoU Research Program’s rapid enrollment from 2018 onward. While the observed calendar-year pattern conflates true practice shifts with cohort-building effects that cannot be fully disentangled, the existence of this temporal structure motivates including the calendar year of first prescription as a confounder. Adding it as an ordered factor strengthened the effect estimates: HRs for both dementia onset and death were larger in magnitude with tighter confidence intervals for death (Table 4 for HRs and Tables 5, 6 and 7 for cumulative risks, risk differences, and restricted mean time lost at different time points). In contrast, the two lifestyle covariates (smoking, alcohol use) and the combined educational attainment and neighborhood-level characteristics affected the propensity score to a smaller extent and did not change the magnitude and significance of estimated treatment effects.

**Table 4:** Cox hazard ratios (metformin vs. sulfonylureas) for dementia and competing death, under unadjusted, overlap, and stabilized ATE weighted models, in the T2D-restricted cohort and under three covariate sensitivity analyses. Point estimate [95% CI].

| **Weighting** | **Outcome** | **Main** | **No presc.-year** | **No lifestyle** | **No education/**<br>**neighborhood** |
|---|---|---|---|---|---|
| Unadjusted | Dementia | 1.01 [0.91, 1.12] | - | - | - |
|  | Death | 0.82 [0.67, 1.00] | - | - | - |
| Overlap | Dementia | 0.92 [0.84, 1.02] | 0.99 [0.90, 1.08] | 0.91 [0.83, 1.01] | 0.92 [0.84, 1.02] |
|  | Death | 0.67 [0.55, 0.81] | 0.82 [0.68, 1.00] | 0.66 [0.55, 0.80] | 0.66 [0.55, 0.80] |
| Stabilized ATE | Dementia | 0.90 [0.82, 1.02] | 0.99 [0.90, 1.09] | 0.90 [0.81, 1.01] | 0.90 [0.82, 1.01] |
|  | Death | 0.61 [0.49, 0.75] | 0.75 [0.61, 0.92] | 0.61 [0.49, 0.75] | 0.61 [0.49, 0.75] |

**Table 5:** Cumulative incidence (%) of dementia and competing death by treatment arm at 5, 10 and 15 years post-baseline, under stabilized ATE weighting, in the T2D-restricted cohort and under three covariate sensitivity analyses. Point estimate [95% CI]; the “Main” column underlies the curves in Figure 7.

| **Horizon** | **Outcome** | **Arm** | **Main** | **No presc.-year** | **No lifestyle** | **No education/<br>neighborhood** |
|---|---|---|---|---|---|---|
| 5 years | Dementia | Metformin | 9.0 [8.5, 9.4] | 9.1 [8.6, 9.5] | 9.0 [8.5, 9.4] | 9.0 [8.5, 9.4] |
|  |  | Sulfonylureas | 10.0 [8.6, 11.3] | 9.5 [8.3, 10.7] | 10.0 [8.6, 11.4] | 10.0 [8.6, 11.3] |
|  | Death | Metformin | 1.3 [1.1, 1.5] | 1.4 [1.2, 1.5] | 1.3 [1.1, 1.5] | 1.3 [1.1, 1.5] |
|  |  | Sulfonylureas | 2.5 [1.7, 3.4] | 2.1 [1.4, 2.8] | 2.5 [1.7, 3.4] | 2.5 [1.7, 3.4] |
| 10 years | Dementia | Metformin | 18.1 [17.3, 18.9] | 18.2 [17.5, 19.0] | 18.1 [17.3, 18.9] | 18.1 [17.3, 18.9] |
|  |  | Sulfonylureas | 20.1 [18.0, 22.3] | 19.3 [17.3, 21.3] | 20.2 [18.2, 22.4] | 20.0 [18.0, 22.2] |
|  | Death | Metformin | 3.4 [3.1, 3.8] | 3.5 [3.2, 3.9] | 3.4 [3.1, 3.8] | 3.4 [3.1, 3.8] |
|  |  | Sulfonylureas | 6.0 [4.7, 7.4] | 5.2 [4.1, 6.5] | 5.9 [4.7, 7.4] | 6.0 [4.7, 7.4] |
| 15 years | Dementia | Metformin | 28.4 [27.1, 29.6] | 28.7 [27.5, 29.9] | 28.4 [27.1, 29.6] | 28.4 [27.2, 29.6] |
|  |  | Sulfonylureas | 30.3 [27.4, 33.1] | 28.9 [26.3, 31.6] | 30.5 [27.6, 33.3] | 30.4 [27.5, 33.2] |
|  | Death | Metformin | 6.1 [5.4, 6.7] | 6.3 [5.6, 7.0] | 6.1 [5.4, 6.7] | 6.1 [5.4, 6.7] |
|  |  | Sulfonylureas | 8.7 [7.1, 10.5] | 7.7 [6.2, 9.3] | 8.7 [7.1, 10.4] | 8.7 [7.0, 10.5] |

**Table 6:**
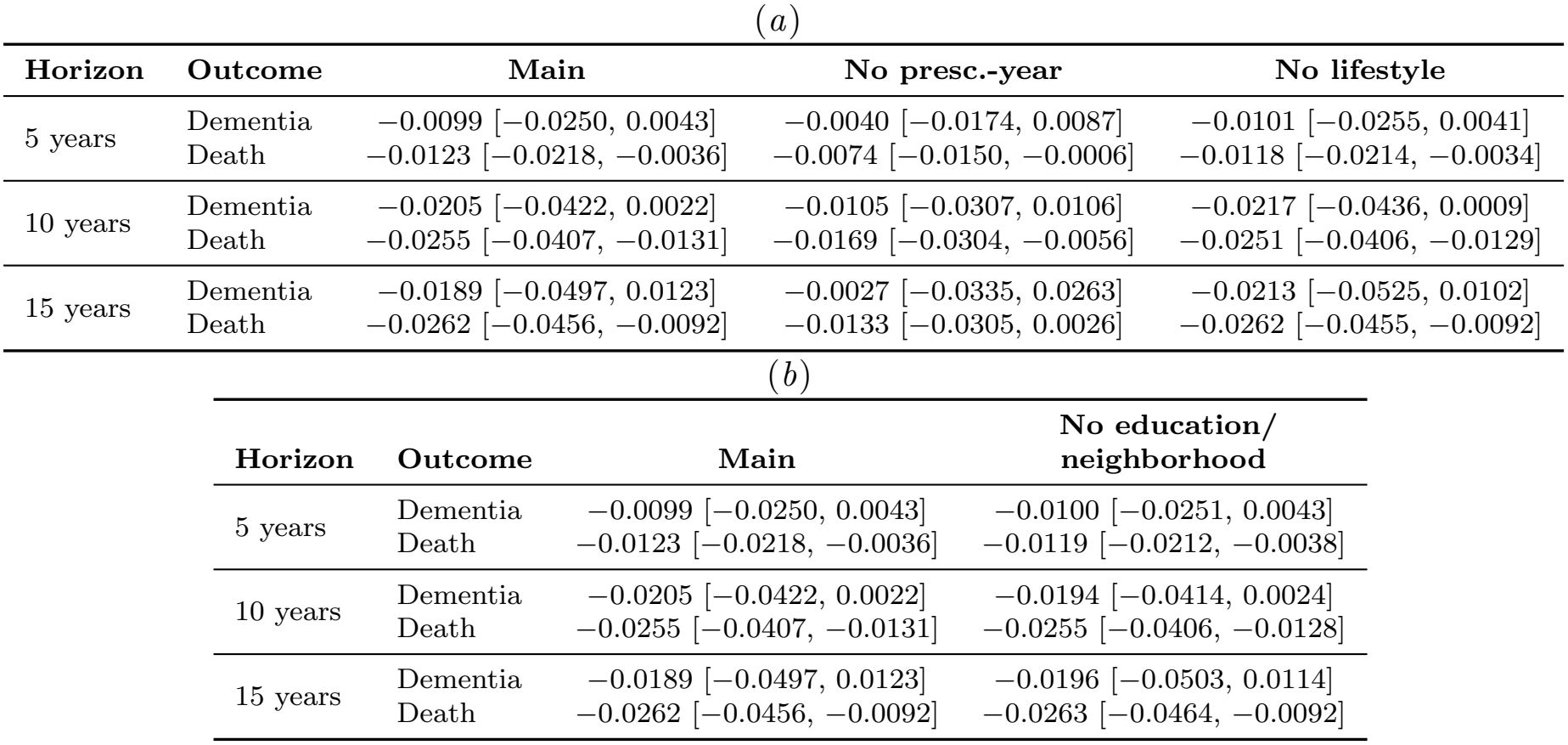
Risk difference (Metformin *−* Sulfonylureas) in cumulative incidence at 5, 10 and 15 years post-baseline, under stabilized ATE weighting, in the T2D-restricted cohort and under three covariate sensitivity analyses. Point estimate [95% CI]; the “Main” column underlies the curves in Figure 8.

| (*a*) |  |  |  |  |
|---|---|---|---|---|
| **Horizon** | **Outcome** | **Main** | **No presc.-year** | **No lifestyle** |
| 5 years | Dementia | −0.0099 [−0.0250, 0.0043] | −0.0040 [−0.0174, 0.0087] | −0.0101 [−0.0255, 0.0041] |
|  | Death | −0.0123 [−0.0218, −0.0036] | −0.0074 [−0.0150, −0.0006] | −0.0118 [−0.0214, −0.0034] |
| 10 years | Dementia | −0.0205 [−0.0422, 0.0022] | −0.0105 [−0.0307, 0.0106] | −0.0217 [−0.0436, 0.0009] |
|  | Death | −0.0255 [−0.0407, −0.0131] | −0.0169 [−0.0304, −0.0056] | −0.0251 [−0.0406, −0.0129] |
| 15 years | Dementia | −0.0189 [−0.0497, 0.0123] | −0.0027 [−0.0335, 0.0263] | −0.0213 [−0.0525, 0.0102] |
|  | Death | −0.0262 [−0.0456, −0.0092] | −0.0133 [−0.0305, 0.0026] | −0.0262 [−0.0455, −0.0092] |
| (*b*) |  |  |  |  |
| **Horizon** | **Outcome** | **Main** | **No education/<br>neighborhood** |  |
| 5 years | Dementia | −0.0099 [−0.0250, 0.0043] | −0.0100 [−0.0251, 0.0043] |  |
|  | Death | −0.0123 [−0.0218, −0.0036] | −0.0119 [−0.0212, −0.0038] |  |
| 10 years | Dementia | −0.0205 [−0.0422, 0.0022] | −0.0194 [−0.0414, 0.0024] |  |
|  | Death | −0.0255 [−0.0407, −0.0131] | −0.0255 [−0.0406, −0.0128] |  |
| 15 years | Dementia | −0.0189 [−0.0497, 0.0123] | −0.0196 [−0.0503, 0.0114] |  |
|  | Death | −0.0262 [−0.0456, −0.0092] | −0.0263 [−0.0464, −0.0092] |  |

**Table 7:** Restricted mean time lost (Metformin *−* Sulfonylureas, in days) at 5, 10 and 15 years post-baseline, under stabilized ATE weighting, in the T2D-restricted cohort and under three covariate sensitivity analyses. Point estimate [95% CI]; the “Main” column underlies the curves in Figure 9.

| **Horizon** | **Outcome** | **Main** | **No presc.-year** | **No lifestyle** | **No education/ neighborhood** |
|---|---|---|---|---|---|
| 5 years | Dementia | −19.0 [−36.8, −2.5] | −10.9 [−26.7, 2.7] | −19.5 [−37.3, −3.1] | −18.4 [−35.7, −2.3] |
|  | Death | −11.8 [−22.9, −3.7] | −7.7 [−16.4, −1.1] | −11.8 [−22.7, −3.7] | −11.4 [−22.0, −3.5] |
| 10 years | Dementia | −40.6 [−84.6, 4.2] | −17.4 [−57.7, 21.7] | −42.4 [−87.3, 2.7] | −40.2 [−83.2, 3.6] |
|  | Death | −44.6 [−74.1, −21.5] | −27.6 [−51.8, −6.8] | −43.8 [−72.8, −20.0] | −43.1 [−71.4, −20.3] |
| 15 years | Dementia | −74.9 [−155.9, 10.7] | −28.2 [−105.3, 47.9] | −79.5 [−162.6, 3.7] | −74.3 [−153.1, 9.8] |
|  | Death | −96.1 [−151.8, −50.8] | −59.4 [−108.2, −19.5] | −95.3 [−149.5, −50.4] | −94.1 [−150.5, −48.6] |

**Figure 10:**
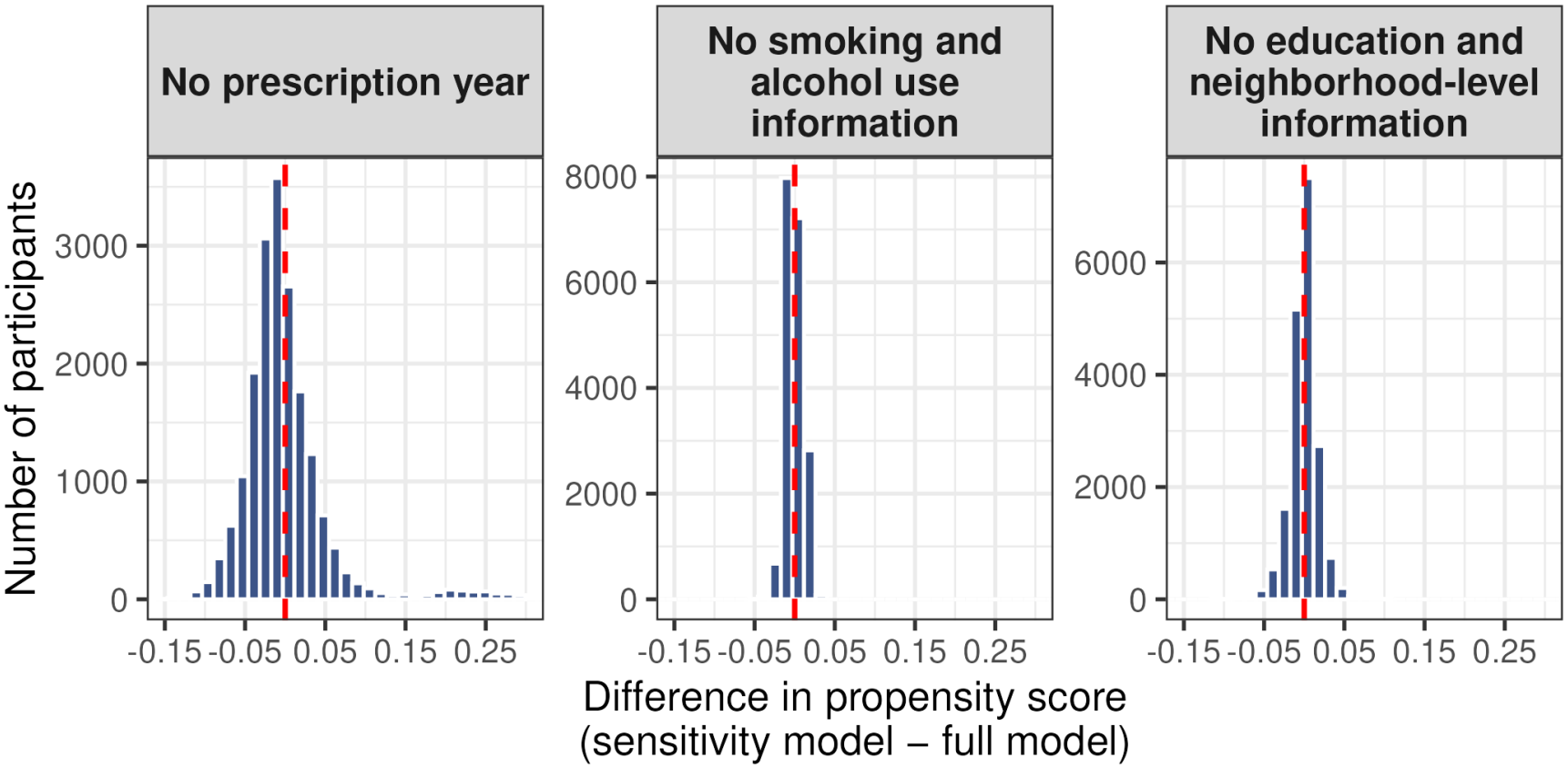
Per-person differences in propensity score under different sensitivity analyses. Dashed red line indicates no difference.

**Figure 11:**
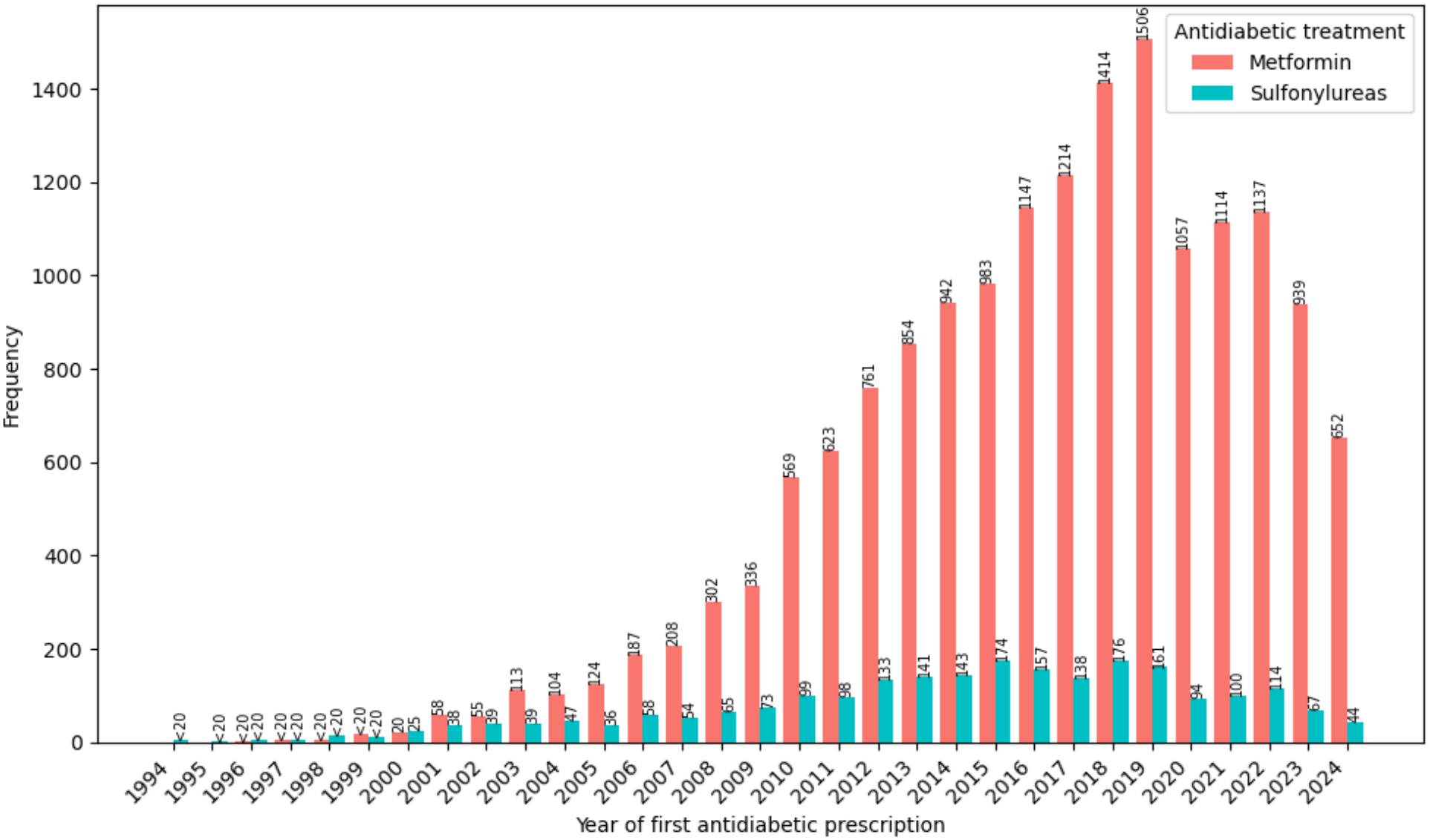
First prescription year counts by trial arm

### T2D ascertainment

Without restricting to participants with a T2D diagnosis on or before baseline, the cohort grows to *N* =29,376 metformin initiators and *N* =3,440 sulfonylurea initiators, with 4,105 dementia events (metformin: 3,500, sulfonylureas: 605) and 1,140 death events (metformin: 912, sulfonylureas: 228); 281 deaths occurred after dementia onset (metformin: 217, sulfonylureas: 64). The full baseline characteristics appear in Tables 8 and 9. Effect estimates were similar to the main (T2D-restricted) analysis (under stabilized ATE weighting, HR dementia: 0.93, [0.85, 1.03]; competing death: 0.63, [0.51, 0.76]). Full unadjusted/overlap HRs for dementia and death and the corresponding cumulative risks, risk differences, and restricted mean time lost at different time points appear in Tables 10, 11, 12 and 13. Restricting the cohort to participants with a T2D diagnosis thus strengthened the dementia signal slightly but left the mortality benefit unchanged.

**Table 8:**
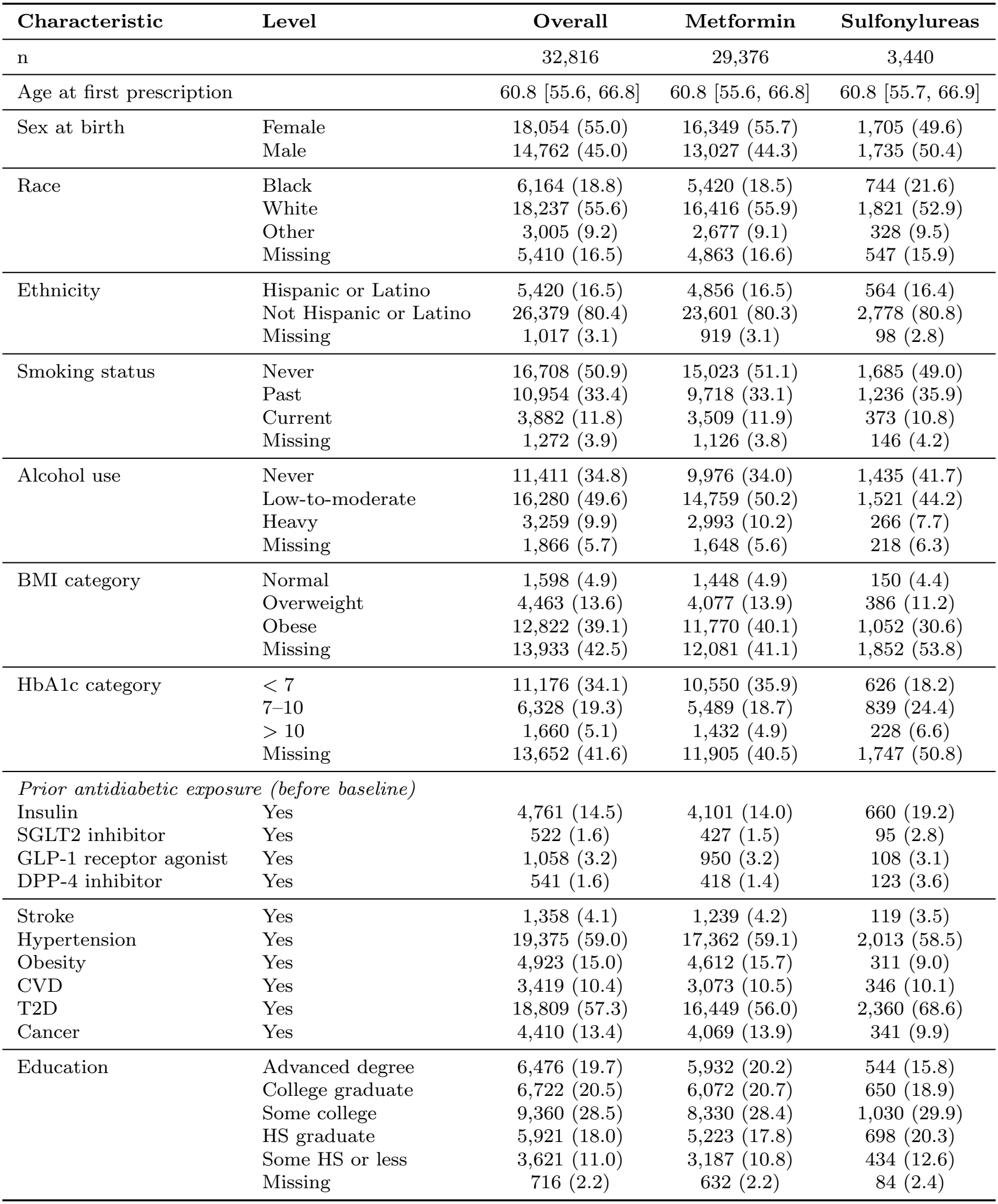
Baseline characteristics of metformin and sulfonylurea initiators including participants without T2D at baseline. Continuous variables: median [interquartile range], categorical variables, n (%).

| **Characteristic** | **Level** | **Overall** | **Metformin** | **Sulfonylureas** |
|---|---|---|---|---|
| n |  | 32,816 | 29,376 | 3,440 |
| Age at first prescription |  | 60.8 [55.6, 66.8] | 60.8 [55.6, 66.8] | 60.8 [55.7, 66.9] |
| Sex at birth | Female | 18,054 (55.0) | 16,349 (55.7) | 1,705 (49.6) |
|  | Male | 14,762 (45.0) | 13,027 (44.3) | 1,735 (50.4) |
| Race | Black | 6,164 (18.8) | 5,420 (18.5) | 744 (21.6) |
|  | White | 18,237 (55.6) | 16,416 (55.9) | 1,821 (52.9) |
|  | Other | 3,005 (9.2) | 2,677 (9.1) | 328 (9.5) |
|  | Missing | 5,410 (16.5) | 4,863 (16.6) | 547 (15.9) |
| Ethnicity | Hispanic or Latino | 5,420 (16.5) | 4,856 (16.5) | 564 (16.4) |
|  | Not Hispanic or Latino | 26,379 (80.4) | 23,601 (80.3) | 2,778 (80.8) |
|  | Missing | 1,017 (3.1) | 919 (3.1) | 98 (2.8) |
| Smoking status | Never | 16,708 (50.9) | 15,023 (51.1) | 1,685 (49.0) |
|  | Past | 10,954 (33.4) | 9,718 (33.1) | 1,236 (35.9) |
|  | Current | 3,882 (11.8) | 3,509 (11.9) | 373 (10.8) |
|  | Missing | 1,272 (3.9) | 1,126 (3.8) | 146 (4.2) |
| Alcohol use | Never | 11,411 (34.8) | 9,976 (34.0) | 1,435 (41.7) |
|  | Low-to-moderate | 16,280 (49.6) | 14,759 (50.2) | 1,521 (44.2) |
|  | Heavy | 3,259 (9.9) | 2,993 (10.2) | 266 (7.7) |
|  | Missing | 1,866 (5.7) | 1,648 (5.6) | 218 (6.3) |
| BMI category | Normal | 1,598 (4.9) | 1,448 (4.9) | 150 (4.4) |
|  | Overweight | 4,463 (13.6) | 4,077 (13.9) | 386 (11.2) |
|  | Obese | 12,822 (39.1) | 11,770 (40.1) | 1,052 (30.6) |
|  | Missing | 13,933 (42.5) | 12,081 (41.1) | 1,852 (53.8) |
| HbA1c category | < 7 | 11,176 (34.1) | 10,550 (35.9) | 626 (18.2) |
|  | 7–10 | 6,328 (19.3) | 5,489 (18.7) | 839 (24.4) |
|  | > 10 | 1,660 (5.1) | 1,432 (4.9) | 228 (6.6) |
|  | Missing | 13,652 (41.6) | 11,905 (40.5) | 1,747 (50.8) |
| *Prior antidiabetic exposure (before baseline)* |  |  |  |  |
| Insulin | Yes | 4,761 (14.5) | 4,101 (14.0) | 660 (19.2) |
| SGLT2 inhibitor | Yes | 522 (1.6) | 427 (1.5) | 95 (2.8) |
| GLP-1 receptor agonist | Yes | 1,058 (3.2) | 950 (3.2) | 108 (3.1) |
| DPP-4 inhibitor | Yes | 541 (1.6) | 418 (1.4) | 123 (3.6) |
| Stroke | Yes | 1,358 (4.1) | 1,239 (4.2) | 119 (3.5) |
| Hypertension | Yes | 19,375 (59.0) | 17,362 (59.1) | 2,013 (58.5) |
| Obesity | Yes | 4,923 (15.0) | 4,612 (15.7) | 311 (9.0) |
| CVD | Yes | 3,419 (10.4) | 3,073 (10.5) | 346 (10.1) |
| T2D | Yes | 18,809 (57.3) | 16,449 (56.0) | 2,360 (68.6) |
| Cancer | Yes | 4,410 (13.4) | 4,069 (13.9) | 341 (9.9) |
| Education | Advanced degree | 6,476 (19.7) | 5,932 (20.2) | 544 (15.8) |
|  | College graduate | 6,722 (20.5) | 6,072 (20.7) | 650 (18.9) |
|  | Some college | 9,360 (28.5) | 8,330 (28.4) | 1,030 (29.9) |
|  | HS graduate | 5,921 (18.0) | 5,223 (17.8) | 698 (20.3) |
|  | Some HS or less | 3,621 (11.0) | 3,187 (10.8) | 434 (12.6) |
|  | Missing | 716 (2.2) | 632 (2.2) | 84 (2.4) |

**Table 9:** Baseline characteristics of metformin and sulfonylurea initiators including participants without T2D at baseline: neighborhood-level variables derived from survey linkages using residential zipcodes. The median is provided, along with the interquartile range. Out of *N* = 32,816 participants, 1,328 had missing neighborhood-level data.

| **Characteristic** | **Overall** | **Metformin** | **Sulfonylureas** |
|---|---|---|---|
| Assisted income, % of zipcode | 13.3 [10.4, 16.6] | 13.3 [10.2, 16.5] | 13.3 [10.5, 17.1] |
| High school education, % of zipcode | 88.8 [85.1, 91.7] | 88.8 [85.1, 91.7] | 88.3 [84.5, 91.4] |
| Median household income, \$ | 61,390 [55,344, 74,163] | 61,390 [55,434, 74,163] | 61,024 [54,436, 74,084] |
| No health insurance, % of zipcode | 8.2 [5.6, 11.5] | 8.2 [5.5, 11.5] | 9.2 [5.7, 11.5] |
| Poverty, % of zipcode | 14.9 [11.7, 18.2] | 14.9 [11.7, 18.1] | 14.9 [11.9, 18.9] |

**Table 10:** Cox hazard ratios (metformin vs. sulfonylureas) for dementia and competing death, under unadjusted, overlap, and stabilized ATE weighted models, in the T2D-restricted cohort and the full unrestricted cohort. Point estimate [95% CI].

| **Weighting** | **Outcome** | **T2D-filtered (main)** | **No T2D filter** |
|---|---|---|---|
| Unadjusted | Dementia | 1.01 [0.91, 1.12] | 1.00 [0.91, 1.08] |
|  | Death | 0.82 [0.67, 1.00] | 0.81 [0.68, 0.97] |
| Overlap | Dementia | 0.92 [0.84, 1.02] | 0.94 [0.87, 1.03] |
|  | Death | 0.67 [0.55, 0.81] | 0.69 [0.58, 0.82] |
| Stabilized ATE | Dementia | 0.90 [0.82, 1.02] | 0.93 [0.85, 1.03] |
|  | Death | 0.61 [0.49, 0.75] | 0.63 [0.51, 0.76] |

**Table 11:**
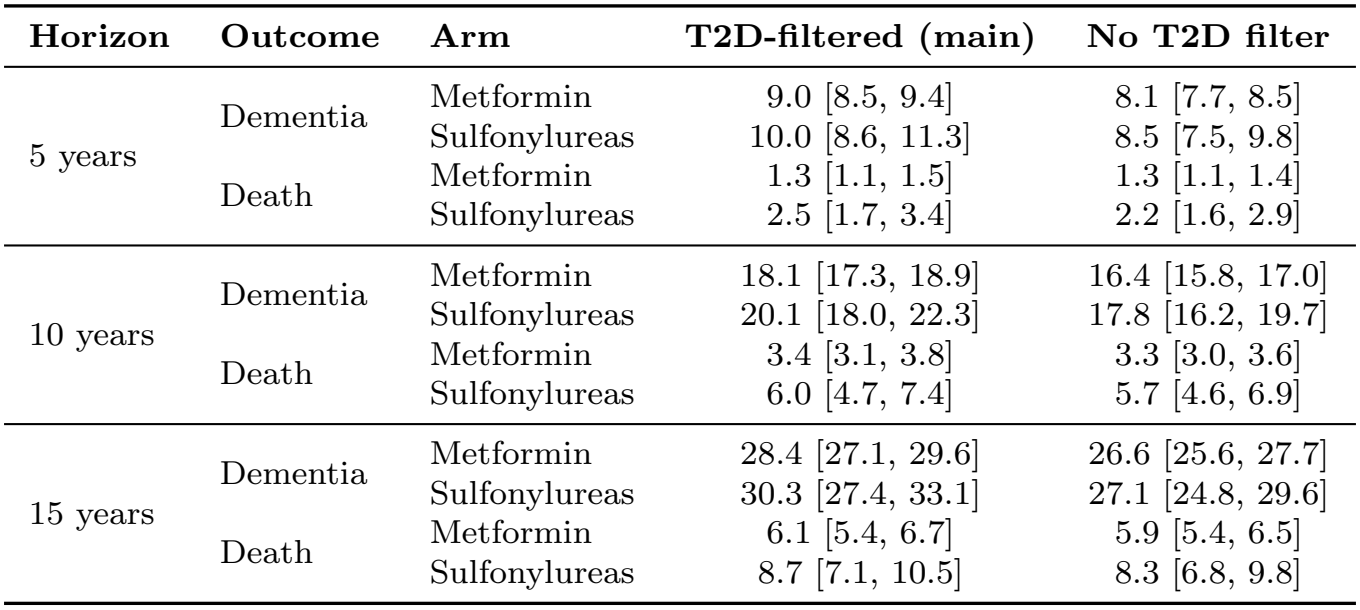
Cumulative incidence (%) of dementia and competing death by treatment arm at 5, 10 and 15 years post-baseline, under stabilized ATE weighting, in the T2D-restricted cohort and the full unrestricted cohort. Point estimate [95% CI].

| **Horizon** | **Outcome** | **Arm** | **T2D-filtered (main)** | **No T2D filter** |
|---|---|---|---|---|
| 5 years | Dementia | Metformin | 9.0 [8.5, 9.4] | 8.1 [7.7, 8.5] |
|  |  | Sulfonylureas | 10.0 [8.6, 11.3] | 8.5 [7.5, 9.8] |
|  | Death | Metformin | 1.3 [1.1, 1.5] | 1.3 [1.1, 1.4] |
|  |  | Sulfonylureas | 2.5 [1.7, 3.4] | 2.2 [1.6, 2.9] |
| 10 years | Dementia | Metformin | 18.1 [17.3, 18.9] | 16.4 [15.8, 17.0] |
|  |  | Sulfonylureas | 20.1 [18.0, 22.3] | 17.8 [16.2, 19.7] |
|  | Death | Metformin | 3.4 [3.1, 3.8] | 3.3 [3.0, 3.6] |
|  |  | Sulfonylureas | 6.0 [4.7, 7.4] | 5.7 [4.6, 6.9] |
| 15 years | Dementia | Metformin | 28.4 [27.1, 29.6] | 26.6 [25.6, 27.7] |
|  |  | Sulfonylureas | 30.3 [27.4, 33.1] | 27.1 [24.8, 29.6] |
|  | Death | Metformin | 6.1 [5.4, 6.7] | 5.9 [5.4, 6.5] |
|  |  | Sulfonylureas | 8.7 [7.1, 10.5] | 8.3 [6.8, 9.8] |

**Table 12:** Risk difference (Metformin *−* Sulfonylureas) in cumulative incidence at 5, 10 and 15 years post-baseline, under stabilized ATE weighting, in the T2D-restricted cohort and the full unrestricted cohort. Point estimate [95% CI].

| **Horizon** | **Outcome** | **T2D-filtered (main)** | **No T2D filter** |
|---|---|---|---|
| 5 years | Dementia | −0.0099 [−0.0250, 0.0043] | −0.0042 [−0.0170, 0.0064] |
|  | Death | −0.0123 [−0.0218, −0.0036] | −0.0089 [−0.0163, −0.0032] |
| 10 years | Dementia | −0.0205 [−0.0422, 0.0022] | −0.0137 [−0.0325, 0.0035] |
|  | Death | −0.0255 [−0.0407, −0.0131] | −0.0236 [−0.0363, −0.0125] |
| 15 years | Dementia | −0.0189 [−0.0497, 0.0123] | −0.0055 [−0.0315, 0.0207] |
|  | Death | −0.0262 [−0.0456, −0.0092] | −0.0233 [−0.0402, −0.0073] |

**Table 13:** Restricted mean time lost (Metformin *−* Sulfonylureas, in days) at 5, 10 and 15 years post-baseline, under stabilized ATE weighting, in the T2D-restricted cohort and the full unrestricted cohort. Point estimate [95% CI].

| **Horizon** | **Outcome** | **T2D-filtered (main)** | **No T2D filter** |
|---|---|---|---|
| 5 years | Dementia | −19.0 [−36.8, −2.5] | −11.5 [−26.3, 0.6] |
|  | Death | −11.8 [−22.9, −3.7] | −9.6 [−17.5, −3.6] |
| 10 years | Dementia | −40.6 [−84.6, 4.2] | −24.1 [−61.6, 9.8] |
|  | Death | −44.6 [−74.1, −21.5] | −37.8 [−60.6, −18.7] |
| 15 years | Dementia | −74.9 [−155.9, 10.7] | −46.4 [−113.7, 15.6] |
|  | Death | −96.1 [−151.8, −50.8] | −84.2 [−129.7, −45.3] |

## 4. Discussion

Our target trial emulation in the NIH AoU database indicates that metformin initiation was robustly associated with a lower risk of death than sulfonylurea initiation in participants aged 50+ at baseline, irrespective of T2D status. Our analysis also suggests a lower risk of dementia onset under metformin initiation, al-though the HR estimate was borderline significant. Results obtained in this large and diverse US cohort corroborate a study conducted in the UK primary care and MGB health systems (Charpignon et al., 2022), as HR estimates closely aligned for dementia onset (UK=0.86, MGB=0.81, AoU=0.90) and death (UK=0.66, MGB=0.57, AoU=0.61). Notably, the beneficial effect of metformin initiation was slightly larger when restricting the population to participants with a coded T2D diagnosis (HR death=0.61 vs. 0.63, HR dementia onset=0.90 vs. 0.93).

Importantly, available AoU data for social and behavioral health variables mentioned as unmeasured confounders in previous work allowed us to test whether estimated propensity scores, HRs, and cumulative risks were sensitive to their inclusion. While all estimates were robust in our study, in part because the baseline distributions of these variables were quite balanced between arms, this observation may not hold in other datasets. In contrast, including the calendar year of first prescription yielded slightly different propensity scores (up to *±*0.25), lower HR and higher risk difference estimates. This finding underscores the importance of accounting for calendar year patterns to reduce residual confounding, especially when the study period spans multiple decades. Indeed, clinical guidelines change over time, influencing physician prescribing preferences. Additionally, mortality rates and dementia incidence rates exhibit both birth cohort effects and secular trends.

Our study has several limitations. (i) The setting (e.g., outpatient vs. inpatient) corresponding to the first prescription was lacking for a sizable proportion of participants. This may result in the mixture of participants who received a prescription to treat chronic T2D with others who experienced a transient hyperglycemic event. (ii) We estimated the intention-to-treat effect of metformin on dementia onset, in the presence of competing death. However, participants are likely to switch to another antidiabetic monotherapy if they experience side effects or to a polytherapy if their T2D deteriorates. Therefore, a per-protocol analysis that accounts for changes in antidiabetic drug exposure and handles time-varying confounders is warranted. (iii) Given the prevalence of missing data, future work could also explore different data imputation schemes. (iv) Our estimates rely on the correct specification of the propensity score model. Future analyses could include dementia and death outcome models and use a doubly-robust approach (e.g., survival targeted maximum likelihood estimation (Benkeser and Hejazi, 2020)) to derive corresponding cumulative risks. (v) The timing of treatment initiation may be inexact, as suggested by a larger-than-expected prevalence of prior insulin prescriptions at baseline. To alleviate this issue, we could learn a latent T2D status or a baseline shift parameter for each participant based on other information in their EHR (e.g., health visits, drug prescriptions, medical exams). Similarly, the diagnosis of dementia is often delayed and only approximate. Our outcome models could be adapted to accommo-date such interval-censored data. Going forward, it would be of interest to replicate our approach in additional health databases (e.g., Kaiser Permanente, FinnGen) to strengthen evidence, assess the repurposing potential of metformin with greater precision, test for effect heterogeneity, and understand underlying mechanisms via mediation analyses.

## Supporting information

ATC codes for antidiabetic medications

ICD codes for cancer (all types; conservative definition)

ICD codes for chronic kidney disease

ICD codes for cardiovascular diseases

ICD codes for Alzheimer's disease and related dementias

ICD codes for hypertension

ICD codes for obesity

ICD codes for stroke (all types)

ICD codes for type 2 diabetes

## Data Availability

This study used data from the NIH All of Us (AoU) Research Program's Controlled Tier Dataset (CDRv9), available to authorized users on the Researcher Workbench. Analysis code is attached as supplemental material.

## Acknowledgments

We gratefully acknowledge All of Us participants, without whom this research would not have been possible, for their contributions. We also thank the National Institutes of Health’s All of Us Research Program for making available the participant data examined in this study.

## Appendix A. Consort diagram

Figure 1 details the cohort-construction flow from the full All of Us Research Program EHR population to the primary T2D-restricted analytic cohort, with perarm participant counts recorded at each filtering step. Of the 481,919 participants with EHR data, 400,846 had no metformin or sulfonylurea prescription and were excluded, leaving 61,209 metformin and 19,864 sulfonylurea arm participants. We then applied the eligibility criteria sequentially: restricting to one drug at initiation and including in each arm only participants whose first recorded prescription corresponded to that arm’s drug removed 10,248 metformin and 13,879 sulfonylurea initiators; requiring age *≥*50 at first prescription removed 18,421 metformin and 1,927 sulfonylurea initiators; excluding a dementia diagnosis at or before baseline removed 1,880 and 156 metformin and sulfonylurea initiators, respectively; excluding a chronic kidney disease (CKD) diagnosis at or before baseline, a metformin contraindication (Imam, 2017), removed 1,162 and 432; and requiring available sex at birth information removed 122 and 30 metformin and sulfonylurea initiators, respectively. These steps yield the unrestricted (“No T2D restriction”) cohort of 29,376 metformin and 3,440 sulfonylurea initiators used in the sensitivity analysis presented in the “T2D ascertainment” paragraph of Section 3 and in Appendix D.4. Finally, restricting to participants with a T2D diagnosis at or before baseline removed 12,927 metformin and 1,080 sulfonylurea initiators, producing the primary cohort of 16,449 metformin and 2,360 sulfonylurea initiators. Within this cohort, 2,422 dementia events and 636 deaths (of which 171 occurred after dementia onset) were observed in the metformin arm, and 478 dementia events and 179 deaths (54 after dementia onset) in the sulfonylurea arm.

## Appendix B. Neighborhood-level socioeconomic variables

The five neighborhood-level covariates are derived by linking each participant’s residential ZIP code (ZIP Code Tabulation Area, ZCTA) to American Community Survey estimates, and describe the participant’s local area rather than the individual socioeconomic information. Each is defined as follows:

- **Assisted income (% of ZIP code):** the percentage of households in the ZIP code receiving public assistance income (e.g., cash publicassistance or Supplemental Nutrition Assistance Program (SNAP)/food-stamp benefits).
- **High-school education (% of ZIP code):** the percentage of adults (aged *≥*25 years) in the ZIP code who have attained at least a high-school diploma or equivalent.
- **Median household income ($):** the median annual household income (in US dollars) across households in the ZIP code.
- **No health insurance (% of ZIP code):** the percentage of the civilian noninstitutionalized population in the ZIP code without any health insurance coverage.
- **Poverty (% of ZIP code):** the percentage of the population in the ZIP code with income below the federal poverty line.

All five measures were standardized to zero mean and unit variance, and missing values were median-imputed, as described in the “Confounders” paragraph of Section 2.

## Appendix C. Baseline characteristics

Baseline characteristics for the primary T2D-restricted cohort are given for categorical variables in Table 2 and for neighborhood-level continuous measures in Table 3. Corresponding tables for the full unrestricted cohort appear in Appendix D.4 (Tables 8 and 9).

## Appendix D. Full results: Figures and Tables

This section includes the full set of figures and tables underlying the results in Section 3. Appendix D.1 presents the main analysis (T2D-restricted, 1,000-bootstrap) balance, propensity score, and outcome plots. Appendix D.2 shows the per-person propensity score differences across sensitivity analyses. Appendix D.3 reports covariate sensitivity analysis effect estimates, and Appendix D.4 a comparison to the cohort with no T2D restriction.

### D.1. Main analysis figures

Figures below are for the primary T2D-restricted cohort under 1,000 bootstrap replicates. Figure 2 shows the estimated propensity score density by arm; Figure 3 shows covariate balance for the mostimbalanced covariates (absolute standardized mean differences *≥* 0.1), including all covariate levels; Figure 7, Figure 8, and Figure 9 show the cumulative incidence function (CIF), risk difference, and restricted-mean-time curves, respectively. The corresponding HRs are reported in Table 1.

### D.2. Sensitivity analysis: propensity score differences

Figure 10 shows the per-person change in estimated propensity score when each covariate group is removed from the model, relative to the main model. Removing the prescription year factor produces the largest shifts. Removing lifestyle or education/neighborhood covariates has a lesser effect.

### D.3. Sensitivity analysis: removing covariates from the propensity score

#### Prescription year shift

As show in Figure 11, incident prescriptions of both drugs rose from a few prescriptions per year before 2000 to a peak of 1,506 metformin and 176 sulfonylurea initiations in 2019 and 2018, respectively, before declining through 2020–2024, consistent with pandemic related disruption to routine primary care. The composition also shifted over time: the metformin-to-sulfonylureas ratio among incident prescriptions rose from roughly 1.5:1 in the early 2000s to approx. 6:1 by the mid-2010s and 10–15:1 by 2019–2024, consistent with clinical guidelines recommending metformin as the first-line antidiabetic therapy. This temporal structure likely also reflects the All of Us Research Program’s recruitment trajectory (enrollment grew rapidly from 2018 onward, drawing on a mix of retrospective and prospective EHR data).

##### Sensitivity analysis results

Dropping prescription year from the propensity score model had the highest effect on the results in our sensitivity analyses. It attenuated the weighted death HR towards the null under both overlap (0.67 *→* 0.82) and stabilized ATE (0.61 *→* 0.75) weighting, and the dementia HR as well (0.90–0.92 *→* 0.99; Table 4). Consistently, it lowered the sulfonylurea-arm death CIF by 10–17% relatively at every horizon (e.g., 2.1% vs. 2.5% at 5 years) while leaving the metformin-arm death CIF unchanged, narrowing the between-arm death contrast. Dementia CIFs in both arms had a lesser effect (Table 5). Death and dementia risk difference and restricted mean time lost were also attenuated when excluding prescription year information (Table 6, Table 7). In contrast, dropping the lifestyle or education/neighborhood covariates leaves all weighted HRs within *∼*0.01 of the main model and arm-specific CIFs within 0.1 percentage points of the main model values at every horizon.

### D.4. Sensitivity analysis: no T2D filtering

To assess whether restricting to participants with a confirmed T2D diagnosis at baseline affects our conclusions, we repeat the full pipeline: baseline characterization, propensity score weighting, and treatment effect estimation on the full eligible cohort of *N* = 32,816 metformin and sulfonylurea initiators, without requiring a T2D diagnosis at or before base-line.

Table 8 reports demographic, clinical, and prior antidiabetic exposure characteristics of this unrestricted cohort by treatment arm, and Table 9 reports the corresponding neighborhood socioeconomic variables. As in the T2D-restricted cohort, metformin and sulfonylurea initiators are similar on most axes. The largest imbalances are concentrated in prior treatment history and metabolic markers. Sulfonylurea initiators more often report prior exposure to insulin (19.2% vs. 14.0%), SGLT2 inhibitors (2.8% vs. 1.5%), and DPP-4 inhibitors (3.6% vs. 1.4%), consistent with sulfonylureas more frequently being used as a later-stage therapy. A larger share of sulfonylurea initiators (68.6%) than metformin initiators (56.0%) carry a formal T2D diagnosis at baseline, reflecting metformin’s broader off-label use in prediabetes and other indications. BMI and HbA1c values are missing more often among sulfonylurea initiators (53.8% and 50.8%, respectively) than metformin initiators (41.1% and 40.5%). Neighborhood-level socioeco-nomic indicators (Table 9) are nearly identical between arms.

Table 10 compares Cox HR for dementia and death between the T2D-restricted (main) and unrestricted cohorts across the three weighting schemes. The direction and approximate magnitude of both associations are preserved when the T2D restriction is lifted: under stabilized ATE weighting, the dementia HR moves from 0.90 [0.82, 1.02] to 0.93 [0.85, 1.03], and the death HR from 0.61 [0.49, 0.75] to 0.63 [0.51, 0.76]. The unadjusted and overlap-weighted models show the same modest, consistent attenuation pattern.

Table 11, Table 12, and Table 13 extend this comparison to cumulative incidence function (CIF), risk difference (RD), and restricted mean time lost (RMST). Relative to the primary T2D-restricted cohort, the full (no T2D filter) cohort shows smaller magnitude RMST and RD contrasts, and lower absolute CIFs for both outcomes and both arms at every horizon. This is expected, since the full cohort dilutes the T2D-specific dementia and mortality risks with participants that did not have a T2D diagnosis at baseline. For RMST, the dementia contrast is attenuated by 38–41% across the 5/10/15-year horizons in the full cohort relative to the T2D-restricted cohort, while the death contrast is less attenuated (12–19%).

